# Comparing performance of deep convolutional neural network with orthopaedic surgeons on identification of total hip prosthesis design from plain radiographs

**DOI:** 10.1101/2020.03.31.20048934

**Authors:** Alireza Borjali, Antonia F. Chen, Hany S. Bedair, Christopher M. Melnic, Orhun K. Muratoglu, Mohammad A. Morid, Kartik M. Varadarajan

**Author notes:** Corresponding author, Address: 55 Fruit Street, GRJ-12-1223, Boston, MA, 02214.

## Abstract

A crucial step in preoperative planning for a revision total hip replacement (THR) surgery is accurate identification of failed implant design, especially if one or more well-fixed/functioning components are to be retained. Manual identification of the implant design from preoperative radiographic images can be time-consuming and inaccurate, which can ultimately lead to increased operating room time, more complex surgery, and increased healthcare costs. No automated system has been developed to accurately and efficiently identify THR implant designs. In this study, we present a novel, fully automatic and interpretable approach to identify the design of nine different THR femoral implants from plain radiographs using deep convolutional neural network (CNN). We also compared the CNN’s performance with three board-certified and fellowship trained orthopaedic surgeons. The CNN achieved on-par accuracy with the orthopaedic surgeons while being significantly faster. The need for additional training data for less distinct designs was also highlighted. Such CNN can be used to automatically identify the design of a failed THR femoral implant preoperatively in just a fraction of a second, saving time and improving identification accuracy.

## 1. INTRODUCTION

One of the key steps in planning a revision joint replacement procedure is identification of failed implants. Particularly, when one or more well-fixed/functioning components are to be retained, accurate implant identification is crucial. Anecdotally, this is accomplished through a combination of reviewing patient records, personal knowledge of different designs, asking manufacturer representatives, consulting with other colleagues, and using image atlases. A 2012 survey of members (n = 605) of American Association of Hip and Knee Surgeons (AAHKS) found that surgeons and staff spent approximately 20-30 minutes for each revision case (41 hours total per year) to identify the implant preoperatively. They reported using five or more methods to identify the implant including radiographs, hospital records, office records, primary procedure notes and implant sheets ^1^. The survey respondents reported that in about 1/10 cases, they could not identify the implant pre-operatively, which then lead to additional requested implants, added surgical time/complexity, increased blood/bone loss, and increased recovery time for patients^1,2^. In another study, the authors calculated the opportunity cost associated with time spent determining the implant design preoperatively based on the AAHKS survey ^2^. Their projection suggested that cumulative surgeon time spent identifying failed implants could reach 133,000 hours in 2030, costing the healthcare system about $27.4 million based on surgeon Medicare reimbursement (2014 coding dollars) rate. Furthermore, the authors projected that by 2030, up to 50,000 cases may not preoperatively identify the failed implant and in about 25,000 cases, the failed components may not be identified intraoperatively ^2^. When the failed components cannot be identified, all components including well-fixed/functioning components need to be unnecessarily removed to avoid issues due to mismatching components of different THR implant designs. This will further increase blood/bone loss and surgery time and complexity.

The use of non-standardized and manual methods for documentation have been cited as a major barrier in accurate device identification ^1^. Important regulatory changes have occurred in an effort to improve implant identification and traceability, including the Food and Drug Administration’s (FDA) rule requiring manufacturers to label medical devices with unique device identifiers (UDIs), establishment of publicly accessible Global Unique Device Identifier Database (GUDID)^2^, and the mandated inclusion of UDI in the patients’ medical records by the Office of the National Coordinator (ONC) for Health Information Technology and Centers for Medicare and Medicaid Services (CMS) ^3,4^. However, as of 2014, over 10 million patients in the United States (US) are living with a hip or knee replacement ^5^, predating these regulatory changes. A large proportion of patients also undergo revision at an institution different from where they had their index procedure (over 40% by 3 years post primary procedure ^6^). Thus, the challenge of accurate device identification remains for many patients. To date, no automated system has been developed to accurately and efficiently identify THR implant designs based on plain radiographs.

We hypothesized that deep learning (DL) based artificial intelligence algorithms could be trained to automatically identify hip implant designs from radiographic images. In recent years, DL methods have been applied to the interpretation of plain film radiographs with high degrees of success for identification and classification of orthopaedic fractures, staging knee osteoarthritis (OA) severity, and detection of aseptic loosening, to name a few ^7-17^. In a previous pilot study, we successfully trained a DL method for the first time to classify a given THR radiograph into one of three possible femoral component designs^18^. Although, this represented a limited task and we did not compare the model’s performance with orthopaedic surgeons, the results were highly encouraging with the model achieving 100% accuracy. The current study is an expansion of our pilot study to investigate the robustness of our proposed DL method on a larger dataset to identify more hip implant designs. It is important to mention that expanding the model to identify more hip implant designs is not as trivial as re-running the same previous model on new data and it requires model modification and entirely new model training process. In particular, we wanted to investigate whether the model performance on the prior simpler task of identifying 3 designs would be matched/maintained by a new model trained to perform a more complex task of identifying several additional designs. Furthermore, in the current study we provided human experts (orthopaeidc surgeons) performance as a baseline to put the proposed model’s value in perspective. On that premise, the goals of the present study were to: 1) train a deep convolutional neural network (CNN) on an expanded dataset involving nine different femoral implant designs, 2) compare model performance to that of orthopaedic surgeons with in regards to accuracy and time spent in performing the task, and 3) visualize the model’s decision making process through the use of saliency maps.

## 2. MATERIALS AND METHODS

We conducted a retrospective study with institutional review board (IRB) approval, using previously collected imaging data at two institutions during 2018. Post-surgery anterior-posterior (AP) hip x-rays of 402 primary THR patients with nine different commonly used THR femoral implant designs were utilized. The x-rays were labeled based on the design of the implant’s femoral stem as recorded in the primary surgery operative note. The stem designs used were: 1) Accolade II (Stryker corporation, Mahwah, NJ, USA), 2) Anthology (Smith & Nephew, London, UK), 3) Corail (DePuy Synthes companies, Warsaw, IN, USA), 4) M/L Taper (Zimmer Biomet, Warsaw, IN, USA), 5) S-ROM, (DePuy Synthes companies, Warsaw, IN, USA), 6) Summit (Johnson & Johnson Medical Devices, New Brunswick, NJ, USA), 7) Taperloc Standard Offset, 8) Taperloc High Offset (Zimmer Biomet, Warsaw, IN, USA), and 9) Versys (Zimmer Biomet, Warsaw, IN, USA). Table 1 demonstrates the THR patient information and the distribution of implant designs. Of note, all Corail and Versys stems in the study had a collar.

**Table 1.** Total hip replacement (THR) patient information

| THR Implant Design | Number of patients | Age (Mean ± Standard Deviation) | Male | Female |
| --- | --- | --- | --- | --- |
| Accolade II | 130 | 62.3 ± 13.0 | 67 (51.5%) | 63(48.5%) |
| Corail | 89 | 66.6 ± 13.1 | 40 (44.9%) | 49 (55.1%) |
| M/L Taper | 31 | 64.2 ± 11.6 | 20 (64.5%) | 11 (35.5%) |
| Summit | 31 | 71.2 ± 10.3 | 7 (22.6%) | 24 (77.4%) |
| Anthology | 26 | 65.5 ± 10.8 | 15 (57.7%) | 11 (42.3%) |
| Versys | 26 | 67.1 ± 10.1 | 10 (38.5%) | 16 (61.5%) |
| S-ROM | 24 | 56.9 ± 16.5 | 16 (66.7%) | 8 (33.3%) |
| Taperloc Standard Offset | 24 | 68.4 ± 11.3 | 12 (50.0%) | 12 (50.0%) |
| Taperloc High Offset | 21 | 62.3 ± 9.1 | 12 (57.1%) | 9 (42.9%) |
| Total | 402 | 64.9 ± 11.7 | 199 (49.5%) | 203 (50.5%) |

Figure 1 illustrates radiographs of each of the nine femoral stems. While the acetabular component throughout the radiographs had similar designs, the stem designs varied more substantially. The medial offset, vertical height, neck-shaft angle, stem length and distal relief of the stems were different. Furthermore, Corail and Versys stems had collars, and the S-ROM stem had a modular metaphyseal sleeve.

**Figure 1.**
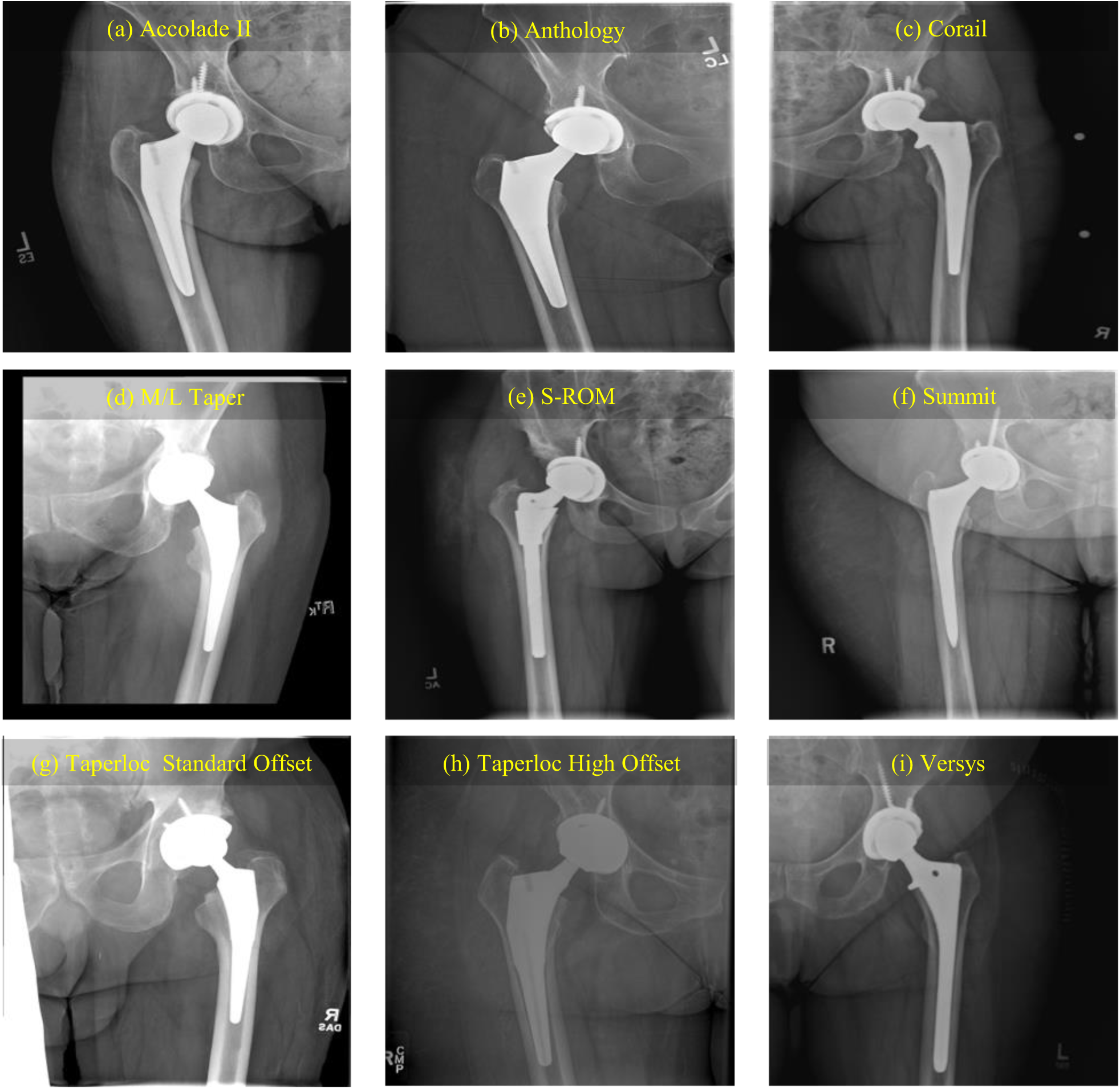
Nine different designs of commonly used total hip replacement (THR) implants: (a) Accolade II, (b) Anthology, (c) Corail, (d) M/L Taper, (e) S-ROM, (f) Summit, (g) Taperloc Standard Offset, (h) Taperloc High Offset, and (i) Versys.

The x-rays were anonymized prior to analysis and the annotations were removed. Split validation method was implemented by randomly dividing the x-rays dataset into training, validation, and final test subsets with 80:10:10 split ratio ^19^. The split ratio was maintained among all the THR implant designs x-rays to make sure that each design had representative x-rays in the training, validation, and test subsets.

The CNN was trained on 320 x-rays in the training subset with online data augmentation. Online data augmentation created new data during the training process by applying random transformations on the base training subset. These random transformations included horizontal flip effectively making the model insensitive to right or left hip x-ray, rotation between 0° and 45°, width and height shift between 0% and 80%, shearing between 0% and 50%, and magnification between 0% and 90%. Since these transformations were applied randomly at each epoch, the CNN saw a different set of x-rays every epoch and it was effectively trained on 320,000 x-rays after 1000 epochs (320 x-rays × 1000 epochs). We chose this heavy data augmentation process since the implant design within each group remained almost constant despite small differences in e.g. size and offset. These implants were all mass-produced off-the-shelf THR implants and all were implanted during the same year; hence the designs were very consistent within each group. Online data augmentation created realistic new x-rays, preserving the same constant design of each THR implant, while compensating for intrinsic x-rays variations. We successfully followed this online data augmentation method in our previous work ^18^ with smaller range (horizontal flip, rotation between 0° and 25°, width and height shift between 0% and 15%, shearing between 0% and 10%, and magnification between 0% and 15%), and less number of epochs (350 epochs). We had to increase both the data augmentation range and number of epochs in this study due to the more complex classification task compared to our previous study (nine THR implant designs in this study vs. three THR implant designs in our previous study). Hyper-parameters including initial learning rate, learning rate decay, regularization, batch size, online data augmentation range, and number of epochs were optimized iteratively on the validation dataset using a grid search strategy. DenseNet-201 ^20^ CNN architecture with 201 layers was used as the base architecture and the final classifier nodes were replaced with nine nodes for categorizing the x-rays into the nine THR implant designs. The CNN was initialized with a random Gaussian distribution weight. CNN’s top-1 and top-3 accuracy were calculated on the test subset, which had not been included in the training or validation process. Top-1 accuracy (conventional accuracy) indicated the percentage of the time that CNN assigned the highest probability (top-1) to the correct THR implant design for each x-ray in the test subset. Top-3 accuracy indicated the percentage of the time that CNN assigned either the highest, second highest, or third highest probability (top-3) to the correct THR implant design for each x-ray in the test subset. While explaining the exact architecture of the CNN is beyond the scope of this paper, we cite the relevant reference for detail ^20^.

Image-specific saliency maps were implemented to visualize the CNN’s decision-making process ^21^. To depict the image-specific saliency map, all the pixels of a given x-ray were ranked based on their relative influence on the CNN’s classification score. Then this ranking was visualized by assigning a color spectrum from red to blue to every pixel of the x-ray based on their level of influence, where red denoted higher relative influence than blue, on the CNN’s classification score. Saliency map demonstrated the most influential pixels on the CNN’s classification score, and in another word, showed where in the x-ray the CNN “looked” at to make a classification. Using saliency maps made the CNN’s results more interpretable and increased the confidence in its output by showing that the CNN considered relevant THR implant features in its decision-making process.

The CNN was trained using Adam optimizer (initial learning rate = 0.001, beta 1 = 0.9, beta 2 = 0.999, epsilon = 1 e-8), with a batch size of 5 for 1000 epochs with early stoppage criteria. The CNN was implemented with Tensorflow (Keras) running on a workstation comprised of an Intel(R) Xeon(R) Gold 6128 processor, 64GB of DDR4 RAM and an NVIDIA Quadro P5000 graphic card. After achieving optimum performance on the validation subset, the CNN was tested on the holdout test subset and the results were reported as the CNN performance.

We compared the CNN’s performance with orthopaedic surgeons’ performance in detecting the THR implant designs. We provided three high-volume board-certified and fellowship trained orthopaedic surgeons with the same information as the CNN. These orthopaedic surgeons were shown the test x-ray images in a blinded fashion without access to any other information about the patients, such as medical history or additional radiographs. The experts were asked to identify each implant following the normal procedure they would follow in their clinical practice when they do not have access to the implant identification record (e.g. use an image atlas, rely on personal experience, send to company representative, google search, etc.). The experts reported the implant type, the method they used to identify each implant, and the time they spent in performing the task. The orthopaedic surgeons’ accuracy and processing time were measured on the entire test subset and were compared with the CNN’s performance.

## 3. RESULTS

The CNN achieved 100% top-1 accuracy in identifying 5 of 9 THR implant designs namely: Accolade II, Corail, S-ROM, Summit, and Versys (Figure 2). Furthermore, CNN achieved 100% top-3 accuracy in identifying 7 of 9 THR implant designs (M/L Taper, Taperloc Standard Offset, Accolade II, Corail, S-ROM, Summit, and Versys; Figure 2). However, for Anthology design, both CNN top-1 and top-3 accuracy were only 33%, and for Taperloc High Offset, the CNN failed to correctly identify the implant on the x-rays of the test subset (top-1 and top-3 accuracy = 0%). We implemented image-specific saliency maps to visualize the CNN’s decision-making process. Figure 3 shows an example x-ray of each THR implant designs and the corresponding CNN saliency map. Colored regions in the saliency map indicated the most influential regions on the CNN performance, where red denoted higher relative influence than blue.

**Figure.2.**
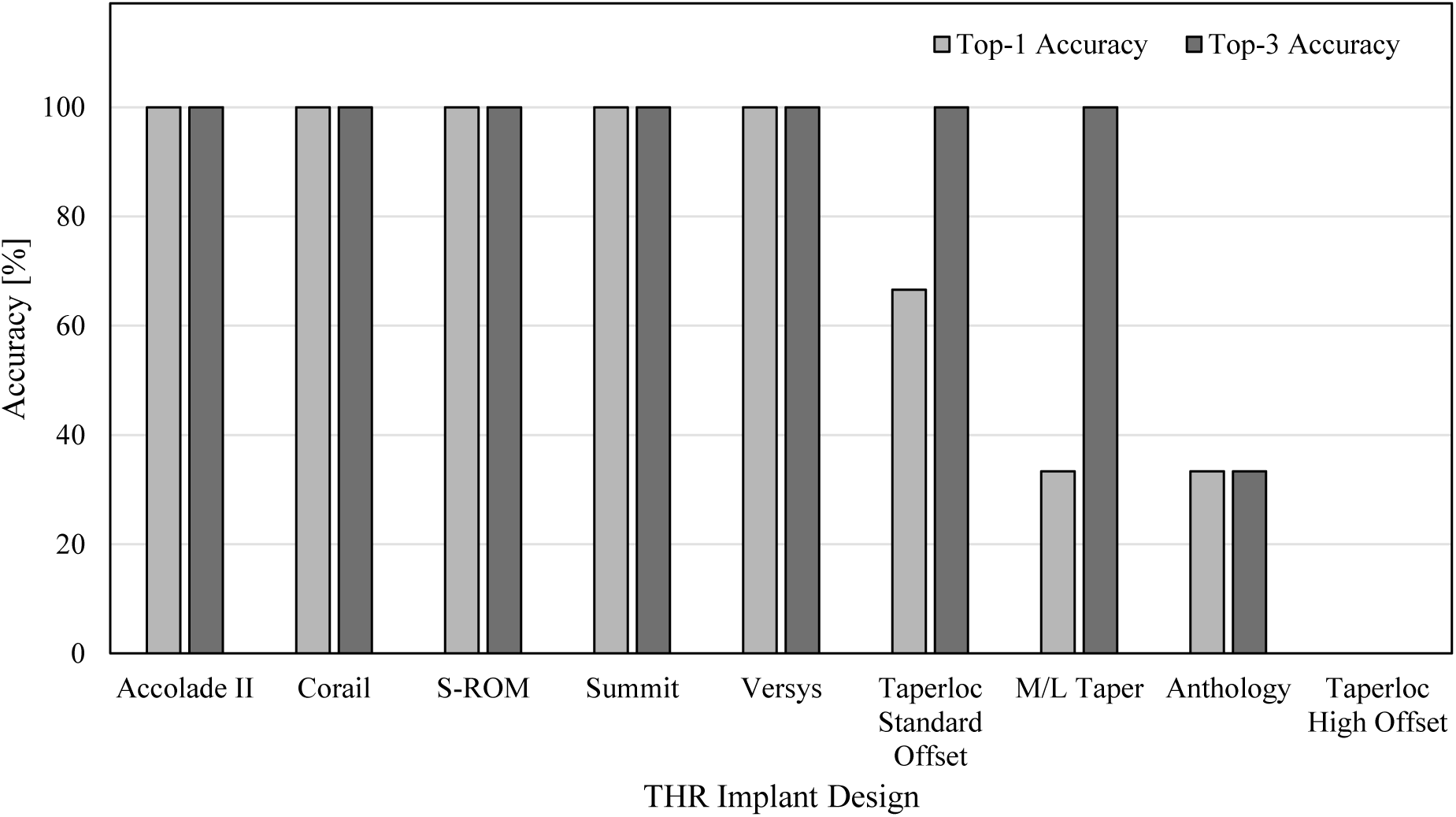
Convolutional neural network (CNN) top-1 and top-3 accuracy identifying nine type of different total hip replacement (THR) femoral implant designs in the test subset.

**Figure 3.**
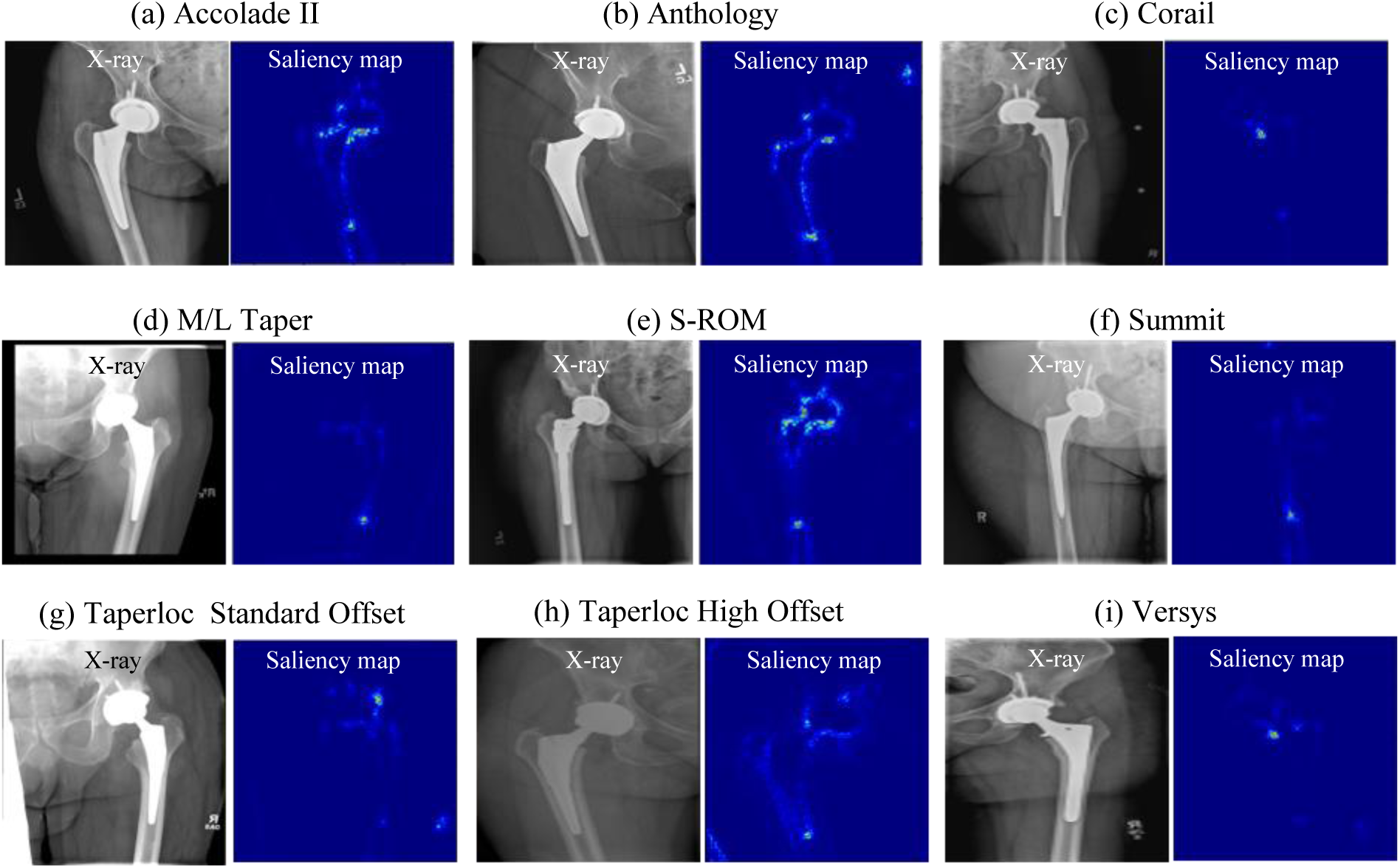
Nine total hip replacement (THR) femoral implant designs and the corresponding convolutional neural network (CNN) saliency maps showing (a) Accolade II, (b) Anthology, (c) Corail, (d) M/L Taper, (e) S-ROM, (f) Summit, (g) Taperloc Standard Offset, (h) Taperloc High Offset, and (i) Versys. Saliency maps indicated most influential regions on the CNN’s performance where red denoted higher relative influence than blue.

Saliency maps illustrated that CNN identified the design of implants by “looking at” the implants while ignoring the background (Figure 3). Figure 4 shows an example x-ray (Accolade II), and the corresponding saliency maps during the training process as a function of training epoch. Number of epochs indicates the number of times that CNN was trained on the entire train subset x-rays. Additionally, this figure demonstrated that the machine gradually focused on the implant’s outline and geometry as the function of the training process and learned to identify the implant. For Corail and Versys THR femoral implant designs that had unique collar features, CNN mainly focused on this feature to identify both designs with 100% top-1 accuracy (Figure 5). On the other hand, the CNN identified Summit THR by only considering the tip of the stem (Figure 6), which was a unique feature of this design.

**Figure 4.**
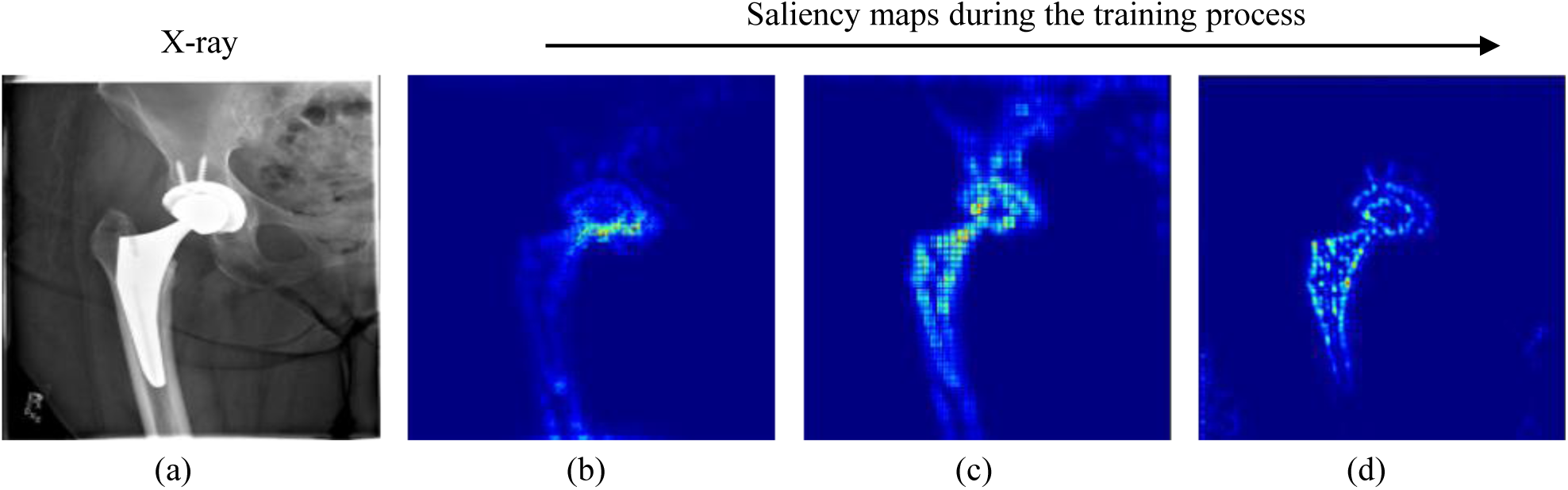
Convolutional neural network (CNN) training process for (a) an example Accolade II x-ray, and the corresponding saliency maps at (b) 1 epoch, (c) 100 epochs, and (d) 500 epochs.

**Figure 5.**
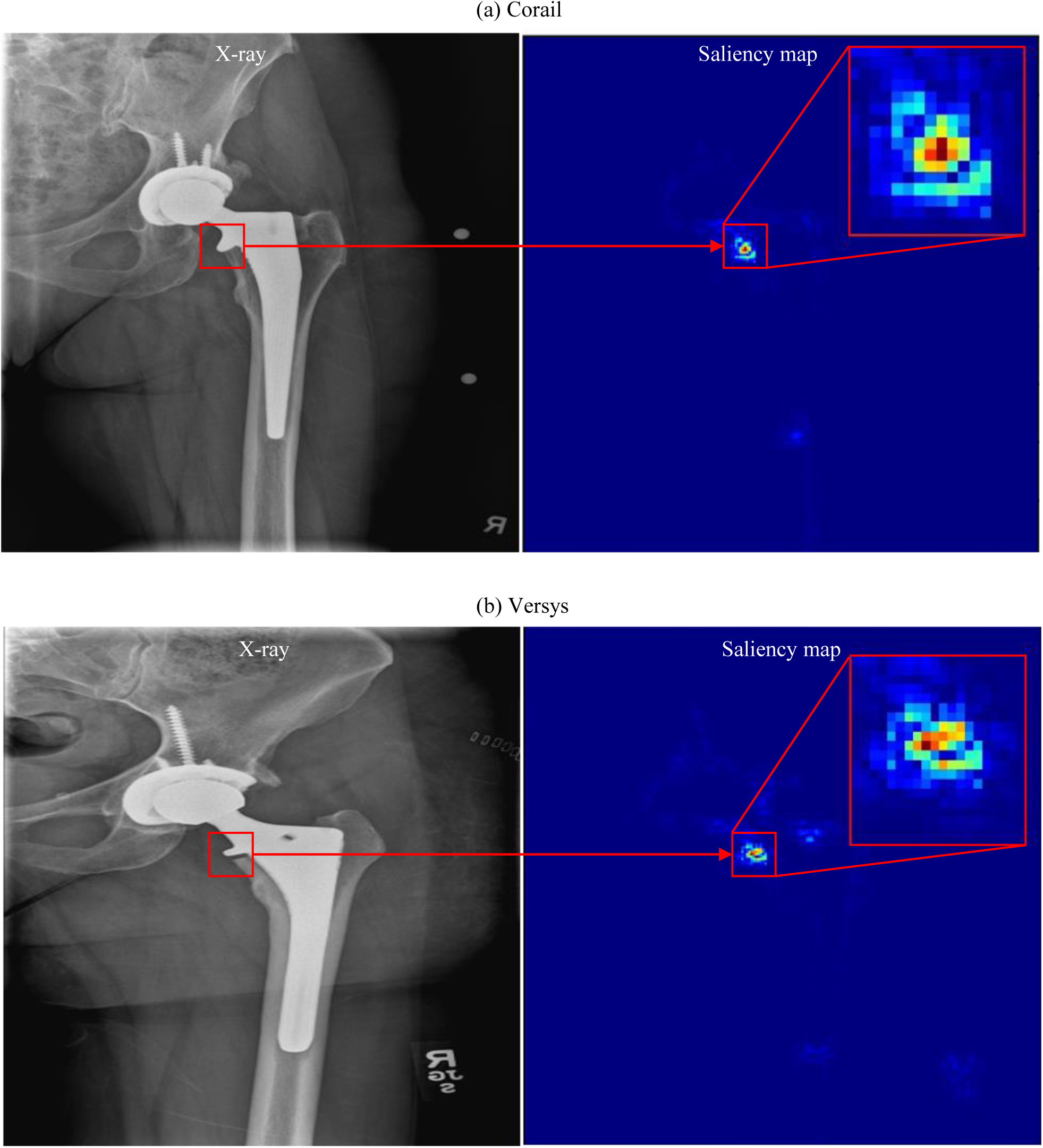
X-ray and corresponding saliency map for (a) Corail and (b) Versys total hip replacement (THR) femoral implants. Saliency maps indicated most influential regions on the convolutional neural network (CNN) performance where red denoted higher relative influence than blue.

**Figure 6.**
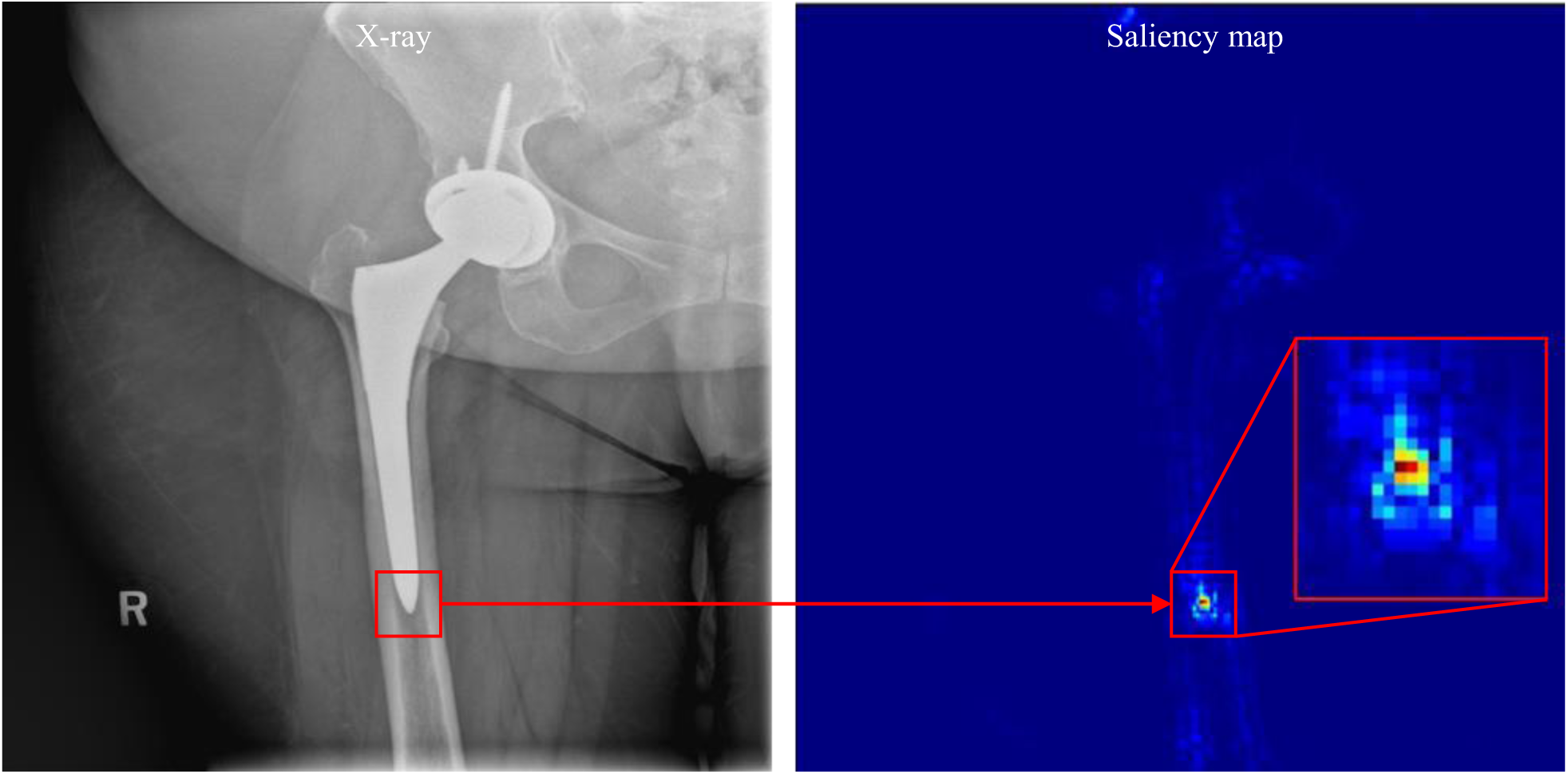
X-ray and corresponding saliency map for Summit total hip replacement (THR) femoral implant. Saliency map indicated the most influential regions on the convolutional neural network (CNN) performance where red denoted higher relative influence than blue.

The tip of Summit THR implant stem had a unique tapered geometry compared to the other designs (Figure 7), which the CNN was able to leverage to accomplish the identification task. For identifying Accolade II and S-ROM THR implant designs, the CNN focused on a combination of the acetabular cup, stem neck, stem tip and the metaphyseal sleeve geometry (for S-ROM) and was able to identify these designs with 100% top-1 accuracy (Figure 3 [a] and Figure 3 [e]). In contrast, the CNN struggled to identify Anthology, M/L Taper, Taperloc Standard Offset, and Taperloc High Offset THR implant designs. In contrast to Versys, Corail, Summit, and S-ROM THR implant designs that had unique distinguishable features that CNN used to identify them, M/L taper, Anthology, Taperloc Standard Offset, and Taperloc High Offset designs had less obviously distinct features. Although the CNN achieved 100% top-3 accuracy identifying M/L taper and Taperloc Standard Offset, its top-3 accuracy remained the same as top-1 accuracy and still struggled to identify Anthology and Taperloc High Offset THR implant designs. Table 2 shows all of the misidentified x-rays by the CNN and their correct THR implant designs.

**Table 2.** Misidentified x-rays by the CNN

| # | Correct THR Design | CNN Top-1 Identification |
| --- | --- | --- |
| 1 | Anthology | Accolade II |
| 2 | Anthology | Taperloc High Offset |
| 3 | M/L Taper | Accolade II |
| 4 | M/L Taper | Accolade II |
| 5 | Taperloc High Offset | M/L Taper |
| 6 | Taperloc High Offset | Accolade II |
| 7 | Taperloc Standard | Taperloc High Offset |

**Figure 7.**
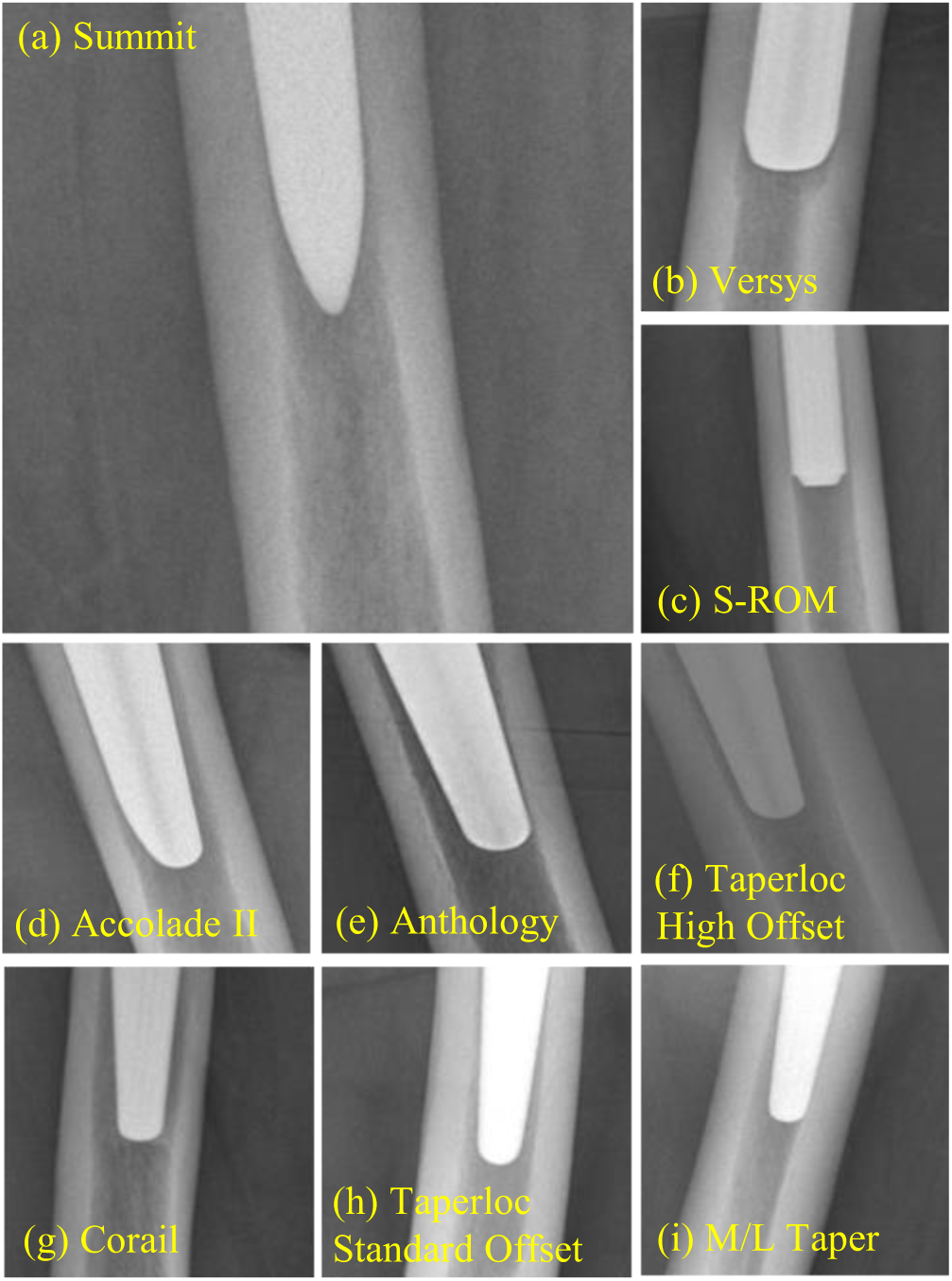
Tip of the stem of nine total hip replacement (THR) implants showing (a) Summit, (b) Versys, (c) S-ROM, (d) Accolade II, (e) Anthology, (f) Taperloc High Offset, (g) Corail, (h), Taperloc Standard Offset, and (i) M/L Taper.

The CNN misidentified Anthology, M/L taper, and Taperloc High Offset THR implants as Accolade II most of the times (4 out of 6). The CNN had the worst performance (0% top-1 and top-3 accuracy) in identifying Taperloc High Offset THR design that had the least overall number of x-rays in the dataset. Furthermore, CNN misidentified Taperloc Standard Offset THR implant as Taperloc High Offset THR implant. The lack of data coupled with higher degree of similarities between designs accounted for the lower performance in these cases.

We also compared the CNN’s performance with that of the orthopaedic surgeons’ on the test subset in detecting the corresponding THR implant designs (Figure 8). For four THR implant designs (Corail, S-ROM, Summit, and Versys), the CNN and all the surgeons achieved 100% accuracy. The CNN, surgeon 1, and surgeon 3 achieved 100% accuracy in identifying Accolade II THR implant design, while surgeon 2 misidentified one of the Accolade II x-rays in the test subset (Figure 8). The CNN achieved similar accuracy as surgeon 2 and surgeon 3 in identifying Taperloc Standard Offset, while surgeon 1 outperformed the rest (CNN, surgeon 2, surgeon 3) and achieved 100% accuracy. The CNN outperformed surgeon 3 and underperformed surgeon 1 and surgeon 2 in identifying M/L taper. The CNN underperformed all the surgeons in identifying Anthology THR implants. Surgeon 1 and surgeon 3 achieved 100% accuracy in identifying Taperloc High Offset THR implant, while both the CNN and surgeon 2 could not identify this THR implant (Figure 8). Overall the CNN achieved the same or higher performance compared to at least one of the surgeons in identifying eight out of nine THR implant designs, and only underperformed all of the surgeons in identifying one THR implant design (Anthology).

**Figure 8.**
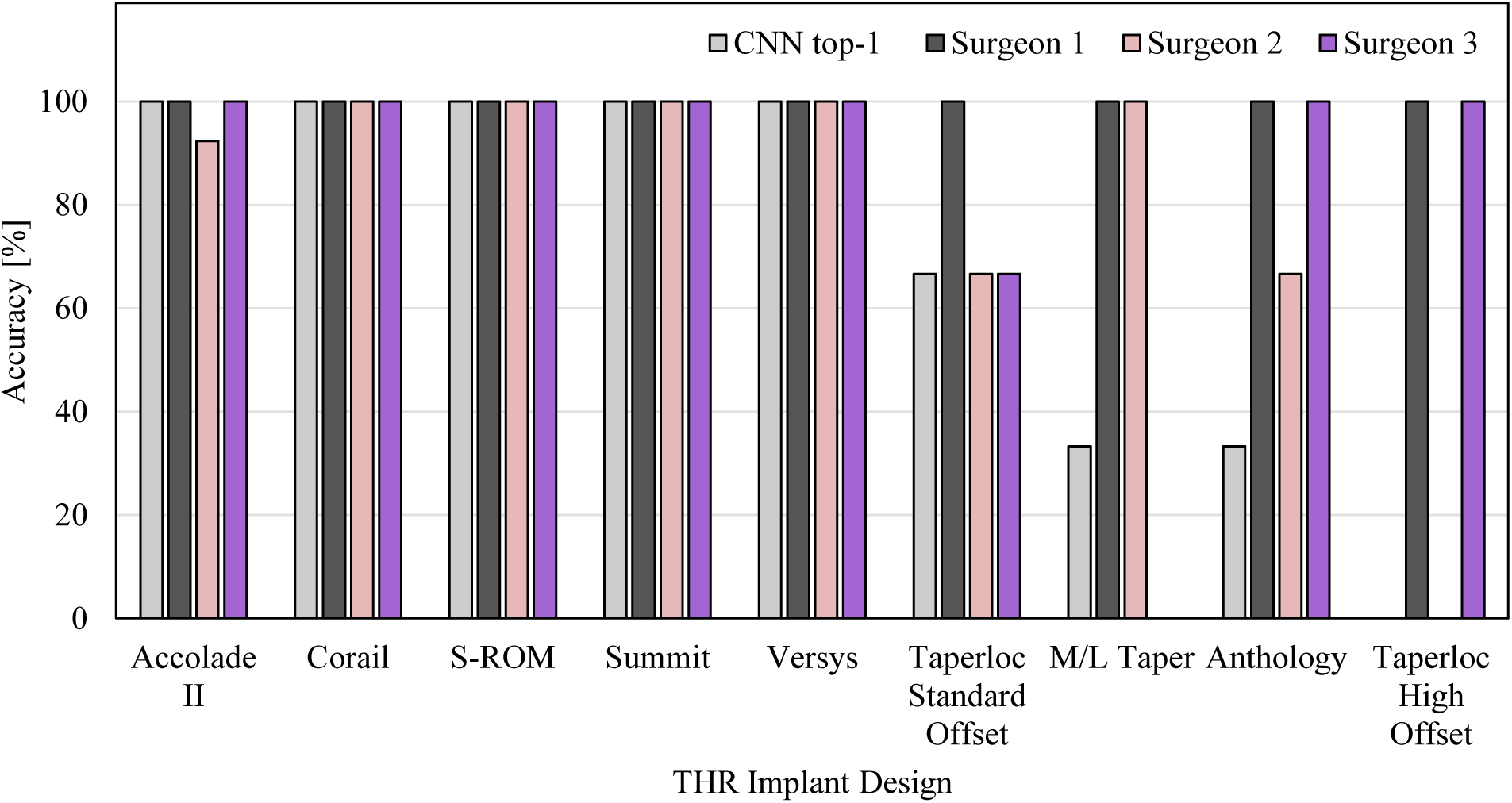
Convolutional neural network (CNN) top-1, surgeon 1, surgeon 2, and surgeon 3 accuracy identifying nine types of different THR implant designs in the test subset

The trained CNN identified the THR implant designs of the test subset (40 x-rays) in 2.4 seconds spending about 0.06 seconds per x-ray. The time spent by the surgeons on the identification task varied based on the identification method. When personal experience was relied upon, the identification time was negligible (perceived as nearly instantaneous). Surgeon 1 used a combination of personal experience (38 x-rays) and online search (2 x-rays; <1 minute per x-ray). Surgeon 2 used a combination of personal experience (29 x-rays), online search (2 x-rays; average 2.5 minutes per x-ray), and consulting with the orthopaedic company representative (9 x-rays; average 15 minutes per x-ray). Surgeon 3 used a combination of personal experience (33 x-rays) and image atlas (7 x-rays; average 3.6 minutes per x-ray). Overall, in cases where surgeons did not identify the THR implant design from personal experience (20/120 x-rays), the three surgeons spent 8.4 minutes on average per x-ray using another identification method (online search, consulting with the orthopaedic company representative, and using image atlas).

## 4 DISCUSSION

In this study, we implemented DL method to automatically detect the design of THR implants from plain film AP radiographs. Other studies have applied DL methods on plain film radiographs for various orthopedic applications ^7–16^. However, to the best of our knowledge, our pilot study identifying three types of THR femoral implant designs^18^, and this study identifying nine types of THR femoral implant designs are the first applications of DL method to automatically detect THR femoral implant designs. We demonstrated that the CNN generally achieved on-par accuracy with the surgeons in identifying these THR implant designs. The CNN achieved the same or higher performance compared with at least one of the surgeons in identifying eight out of nine THR implant designs and underperformed all of the surgeons in identifying one THR implant design (Anthology).

We used online data augmentation combined with high number of epochs to give the CNN ample inputs and opportunity to learn the THR implant designs features. The CNN achieved 100% top-1 accuracy in identifying 5 out of 9 THR implant designs namely: Accolade II, Corail, S-ROM, Summit, and Versys. This further corroborated out pilot study result where the CNN also achieved 100% top-1 accuracy in identifying 3 types of the same THR implant designs (Accolade II, Corail, and S-ROM) ^18^ as this study. The fact that DL did not lose its prior performance in identifying these 3 models was very encouraging. This proved the robustness and scalability of the proposed DL method in analyzing new datasets given appropriate training. Although the CNN did not achieve 100% top-1 accuracy in identifying 4 out of 9 THR implant designs (M/L Taper, Taperloc Standard Offset, Taperloc High Offset, and Anthology), its performance was on-par with orthopaedic surgeons. The orthopaedic surgeons’ accuracy also generally dropped in identifying the same 4 types of THR implant designs. This showed that these 4 THR implant designs were more challenging to identify as they lacked unique features and the design differences were subtler. Furthermore, the number of these THR implant designs x-rays were comparatively low in the dataset that despite heavy data augmentation, the CNN still could not learn to identify them.

In terms of time spent performing the implant identification time, the trained CNN accomplished this in ∼0.06 seconds per x-ray. The surgeon’s identification time varied based on the method utilized. When using their personal experience to identify the THR implant design, this was accomplished in negligible time. However, it increased to an average of 8.4 minutes per x-ray when they used another identification method (online search, consulting with the orthopaedic company representative, and using image atlas), which occurred in about 17% of cases in test subset. It should be mentioned that the surgeons in our study had easy access to all the x-rays in one folder. The time that would had taken them to find the patient’s record, go through the record and identify the relevant imaging data, accesses the file, and finally open the correct x-ray view, which can significantly increase the overall identification time, was not considered. Furthermore, all the THR femoral implant designs in our dataset were more recent designs implanted in 2018. This most likely aided the surgeons with the identification process. We believe these two factors were most likely the reason why the surgeons in our study reported less time identifying the THR implant designs when compared to the 20-30 minutes reported in literature^1^. This further illustrates the importance of the CNN efficient process for preoperative implant identification.

The study result supported our initial hypothesis that a CNN can be trained to provide automated identification of THR femoral implant designs from radiographic images with on-par orthopaedic surgeons’ accuracy while saving significant time and effort. This shows the potential of integrating such CNN in orthopaedic care for identification of failed femoral implants prior to revision procedures. This can potentially have a significant impact on improving patient’s health and decreasing overall cost of revision THR procedures by identifying the right components to replace and saving surgeons significant time to engage in more patient-centric care.

We also used saliency maps to visualize the decision-making process of the CNN and shed light on its process. We showed that CNN looked at each implant’s unique features to make a classification and ignored the background without being explicitly programmed to do so. Saliency maps also revealed that such algorithms may approach a given task different than humans might; for example, using stem tip as an identifying feature, which is not obvious focus point for human reviewers. This is a simple example of how AI may help to obtain new insights on a given task. While a few other studies ^10,11^ have used saliency maps to visualize the CNN’s final output for orthopaedic applications, the use of saliency maps as a tool for pointing out new features, as presented here, is novel.

The primary limitation of this study was the size of the dataset. The CNN’s accuracy was limited for the implants with the least number of x-rays in the dataset, which had subtler geometric differences compared to the other designs. In future studies and with a larger dataset, we can improve the CNN’s performance and consider more THR implant designs. Another limitation of this study was that we only used one AP x-ray per patient, as opposed to clinical practice, where the clinician would consider additional image views to identify the design of a THR implant. In ongoing work, we are incorporating multi-view capabilities to our CNN model. In this study, we presented a novel, fully automatic and interpretable approach to identify the design of nine different THR femoral implants from AP hip x-ray using deep CNN. We compared the CNN’s performance with three board-certified and fellowship trained orthopaedic surgeons and demonstrated that CNN achieved comparable accuracy with the orthopaedic surgeons while being significantly faster. We intend to further develop this AI method to increase its accuracy and add more THR implants designs as well as other implants (e.g. total knee replacement implants). Automated and accurate identification of failed implant components may prove to be a valuable addition to the toolbox of revision arthroplasty surgeons by saving significant time and effort, and reducing likelihood of incorrect pre-operative device identification.

## Data Availability

The IRB protocol does not allow for publicly sharing the data. Specific requests could be directed to the corresponding author.

## AKNOWLEDGEMENT

This study was supported by laboratory sundry funds, and no external funding was received. The authors have no financial disclosures relevant to this study.

## AUTHOR CONTRIBUTIONS

AB, AM, and KV designed the study. AB performed the study. AB, KV, AC, HB, CM, and OM performed the analysis and interpretation of the results. Each author has contributed to the writing and revising the manuscript. All authors have read and approved the final submitted manuscript.

## REFERENCES

1. Wilson, N. A., Jehn, M., York, S. & Davis, C. M. Revision total hip and knee arthroplasty implant identification: Implications for use of unique device identification 2012 AAHKS member survey results. J. Arthroplasty 29, 251–255 (2014).

2. Wilson, N., Broatch, J., Jehn, M. & Davis, C. National projections of time, cost and failure in implantable device identification: Consideration of unique device identification use. Healthcare 3, 196–201 (2015).

3. Weissman, J. S., Krupka, D. C., Zerhouni, Y., Landman, A. & Wilson, N. Transmitting the UDI from the Point of Use to Insurance Claims: Changes in Workflows and Information Systems. (2017).

4. Home | Ortho-tag. Available at: https://www.ortho-tag.com/. x(Accessed: 30th March 2020)

5. Kremers, H. M. et al. Prevalence of total hip and knee replacement in the United States. J. Bone Jt. Surg. - Am. Vol. 97, 1386–1397 (2014).

6. Dy, C. J. et al. Is changing hospitals for revision total joint arthroplasty associated with more complications? Clin. Orthop. Relat. Res. 472, 2006–2015 (2014).

7. Yi, P. H. et al. Deep Learning-Based Identification Of Traditional Hip, Knee, and Shoulder Arthroplasty and Application to Alternative Arthroplasty Designs. in Machine Learning for Healthcare 2018; Stanford, CA2018 (2018).

8. Thomas, K. et al. Automated Classification of Knee X-rays Using Deep Neural Networks Outperforms Radiologist. in Orhopaedic research society (2019). https://www.ors.org/Transactions/65/1807.pdf

9. Krogue JD, Cheng K, Hwang K, et al. Automatic Hip Fracture Identification and Functional Classification with Deep Learning. in Orthopaedic research society (2019). https://www.ors.org/Transactions/65/0108.pdf

10. Tiulpin, A., Thevenot, J., Rahtu, E., Lehenkari, P. & Saarakkala, S. Automatic knee osteoarthritis diagnosis from plain radiographs: A deep learning-based approach. Sci. Rep. 8, 1727 (2018).

11. Olczak, J. et al. Artificial intelligence for analyzing orthopedic trauma radiographs: Deep learning algorithms—are they on par with humans for diagnosing fractures? Acta Orthop. 88, 581–586 (2017).

12. Kitamura, G., Chung, C. Y. & Moore, B. E. Ankle Fracture Detection Utilizing a Convolutional Neural Network Ensemble Implemented with a Small Sample, De Novo Training, and Multiview Incorporation. J. Digit. Imaging (2019). doi:10.1007/s10278-018-0167-7

13. Chung, S. W. et al. Automated detection and classification of the proximal humerus fracture by using deep learning algorithm. Acta Orthop. 89, 468–473 (2018).

14. Rayan, J. C., Reddy, N., Kan, J. H., Zhang, W. & Annapragada, A. Binomial Classification of Pediatric Elbow Fractures Using a Deep Learning Multiview Approach Emulating Radiologist Decision Making. Radiol. Artif. Intell. 1, e180015 (2019).

15. Lindsey, R. et al. Deep neural network improves fracture detection by clinicians. Proc. Natl. Acad. Sci. 115, 11591–11596 (2018).

16. Borjali, A., Chen, A. F., Muratoglu, O. K., Morid, M. A. & Varadarajan, K. M. Detecting mechanical loosening of total hip replacement implant from plain radiograph using deep convolutional neural network. (2019). 1912.00943 [eess.IV]

17. Borjali, A., Chen, A., Muratoglu, O. & Varadarajan, K. M. Detecting Mechanical Loosening Of Total Hip Arthroplasty Using Deep Convolutional Neural Network. Orthop. Proc. 102-B, 133 (2020).

18. Borjali, A., Chen, A. F., Muratoglu, O. K., Morid, M. A. & Varadarajan, K. M. Detecting total hip replacement prosthesis design on plain radiographs using deep convolutional neural network. J. Orthop. Res. jor.24617 (2020). doi:10.1002/jor.24617

19. Borjali, A., Monson, K. & Raeymaekers, B. Predicting the polyethylene wear rate in pin-on-disc experiments in the context of prosthetic hip implants: Deriving a data-driven model using machine learning methods. Tribol. Int. 133, 101–110 (2019).

20. Huang, G., Liu, Z., Van Der Maaten, L. & Weinberger, K. Q. Densely connected convolutional networks. in Proceedings - 30th IEEE Conference on Computer Vision and Pattern Recognition, CVPR 2017 2017-Janua, 2261–2269 (2017).

21. Simonyan, K., Vedaldi, A. & Zisserman, A. Deep Inside Convolutional Networks: Visualising Image Classification Models and Saliency Maps. (2013). 1312.6034 [cs.CV]

